# Characterizing social behavior relevant for tuberculosis transmission in four low- and middle-income countries

**DOI:** 10.1101/2025.10.27.25338900

**Authors:** Kristin N Nelson, Machi Shiiba, Rajan Srinivasan, Tyler S. Brown, Leonardo Martinez, Charlotte Doran, Samanta Biswas, Venkata Raghava, Charfudin Sacoor, Momin Kazi, Herberth Maldonado, Saad Omer, Ben Lopman

**Author notes:** **Corresponding author**: Kristin Nelson, Department of Epidemiology, Rollins School of Public Health, Emory University, 1518 Clifton Road, Atlanta, GA 30322.

## Abstract

**Background:** Tuberculosis (TB) is caused by *Mycobacterium tuberculosis* (*Mtb*), a bacterium which is transmitted through the air. Close, sustained contact can lead to transmission of *Mtb*, but evidence also shows that transmission occurs also in community settings through more transient contact. However, social patterns that influence *Mtb* transmission, and locations that are most central to spread, are likely different by setting.

**Methods:** We use data from the GlobalMix study, which characterized social behavior across four countries, to evaluate differences in age-sex patterning and locations of contact across four moderate- to high-TB burden countries. Healthy individuals self-completed a survey about their daily person-to-person interactions and locations in which they spent time. To capture the *Mtb* exposure profile of participants, we calculated daily exposure-hours from household contacts, close non-household contacts, and casual contacts, stratified by contact age and sex, and weighted by country-, age-, and sex-specific tuberculosis incidence estimates.

**Results:** The most prominent shifts in the profile of exposure occur at entry to primary school (5-9 years) and early adulthood (20-29 years). Community locations varied in their relative importance as locations of transmission by country and age group, with school most important in India and Guatemala, transit more important in Pakistan, and others’ homes most important in Mozambique.

**Conclusions:** Our findings demonstrate that locations of community transmission are likely varied across settings, underscoring the importance of interventions designed specifically for the communities in which they will be implemented.

## Background

Tuberculosis (TB) is the leading infectious cause of death globally from a single pathogen. In 2023, there were an estimated 10.5 million cases and 1.3 million deaths from TB; the vast majority occur in low- and middle-income countries. [1] New vaccines, shorter drug regimens, improved diagnostics, and innovation in active case-finding campaigns, which seek to identify tuberculosis cases outside of traditional healthcare settings, are all promising strategies to tackle entrenched tuberculosis epidemics. However, wielding these tools effectively is essential to maximizing their impact. Understanding the social and cultural forces driving differences in tuberculosis epidemics can lead to a clearer understanding of the interventions that will be useful in different contexts to curb tuberculosis transmission.

TB is caused by *Mycobacterium tuberculosis* (*Mtb*), a bacterium which is transmitted through the air. Close, sustained contact, such as among household members, can lead to transmission of *Mtb*. However, the relative importance of close contacts in transmission overall remains uncertain. [2], [3] Yields from contact investigations, which systematically investigate people who have close interactions with infectious tuberculosis cases to screen them for TB, are generally low: a recent systematic review of studies from low- and middle-income settings reported that only 3-4 % of close contacts were found to have prevalent TB. [4], [5] Indeed, studies which have combined *Mtb* molecular data with information on contacts have concluded that a minority of tuberculosis cases (5-20%) can be attributed to close interactions between tuberculosis cases and household or community members. [6], [7], [8], [9] Meanwhile, evidence continues to accumulate that brief and low-intensity casual contacts (e.g., sitting near someone on a bus or in a church) may be sufficient to spread *Mtb* [10] and modeling studies have further suggested these interactions could in fact be responsible for a substantial portion of transmission. [11] Understanding where these interactions occur can inform siting of community-based interventions, such as educational campaigns, active case finding efforts, or environmental controls (i.e., increasing ventilation) in places where *Mtb* transmission is likely to occur.

Social behaviors, and therefore exposure to and burden of TB, are strongly patterned by age and sex. For example, the infectious form of pulmonary tuberculosis is more common among young adults than among children or older adults, with smear-positive, pulmonary tuberculosis peaking among 20 to 29 year-olds in most high-burden countries.[12] Men are 2.5 times more likely to have smear-positive pulmonary tuberculosis than women, and are less likely to seek and access tuberculosis care.[13] Studies of contact patterns in South Africa and Zambia have suggested that most tuberculosis infections are attributable to transmission from adult men.[14] However, these patterns are likely to differ across countries with unique social, cultural, and religious norms. Identifying demographic groups most likely to contribute to new *Mtb* infections can allow for the careful design of interventions to effectively reach these populations.

Dynamic mathematical models of transmission are useful tools for projecting the impact of interventions. Typically, these models rely on suboptimal information about contact patterns. Seminal studies of social contact were conducted in Western Europe in the early 2000s, distant in time and place from the regions where tuberculosis most common. [15] However, many tuberculosis models continue to rely on these data to simulate the spread of infection in high tuberculosis burden settings in Asia and Africa, which may be insufficient to account for important social and cultural differences between high-income and low-income countries and changes over time in contact patterns. [16], [17], [18] Moreover, early contact studies captured ‘close’ interactions, which involve a conversation or physical touch between two people, but did not capture locations at which individuals spent time where many people share air. [15] As a result, these data may not reflect contact patterns relevant for the spread of an airborne pathogen such as *Mtb* since they emphasize physical contact, which alone is unlikely to spread *Mtb,* and fail to include more ‘casual’ contacts that may. New data from low- and middle countries, which captures these casual interactions in addition to close contacts, offer a more accurate representation of the behaviors that drive transmission.

Previous studies which have examined social contact patterns relevant for *Mtb* transmission did not include children [14] and have focused on countries in southern Africa.[19] They have not evaluated and compared settings across global regions to understand heterogeneity on social behaviors that drive transmission. In this study, we used data from the GlobalMix study to evaluate differences in age-sex patterning and locations of contact across four moderate- to high-TB burden countries and examine their implications for interventions against tuberculosis in diverse settings.

## Methods

### Data collection procedures

The GlobalMix study characterized social behavior across four countries: Pakistan, Mozambique, Guatemala, and India. The study design is described in detail elsewhere [20] and involved a qualitative data collection phase, which included focus group discussions and cognitive interviews, with the objective of adapting the data collection instruments to each site. Households within study sites were enumerated and visited sequentially. In all countries, we defined a household as a group of people who share space for cooking. Within each household, one household member was selected at random and offered participation in the study. Participants completed an enrollment questionnaire which included information on their age, gender, current and past schooling, occupation, and household characteristics.

Field workers trained participants to self-complete a survey about their daily person-to-person interactions and places in which they had spent time. Similar to previous studies, we defined close contact as a two-way conversation with three or more words exchanged in the physical presence of another person. Participants were randomly assigned a day of the week to start reporting for two consecutive days. Participants reported characteristics of contacts including contact age and sex, as well as the duration (<5 minutes, 5-15 minutes, 16-30 minutes, 31 minutes-1 hour, 1-4 hours, and 4+ hours), location (home store / market, school, work, place of worship, etc. and whether this place was indoors or outdoors), and whether physical touch was involved.

To capture community locations in which no close contact may have occurred but where the participant may have shared air with others, we also recorded all locations visited by participants over the two days, the amount of time spent there, and estimated the number of people present. For adults or children unable to complete the diary themselves, field staff assisted in selecting another person (a ‘shadow’) to record the participant’s activities.

### Statistical analyses

We define three types of contact for our analyses: (1) household contacts (contacts with members of a participant’s household); (2) close, non-household contacts (contacts with persons who do not belong to an individual’s household), and (3) people who spent time in the same community location as the participant but did not meet our definition of contact, henceforth ‘casual’ contacts.

We calculated daily exposure-hours by summing the length of each contact using the midpoint of the duration categories (e.g., 2.5 minutes for <5 minutes) and using 4 hours for contacts reported as longer than four hours. Because we did not have information on the age and sex of people present in community locations reported by participants, we based this on the age and sex distributions of study participants who reported visiting the same location type and applied post-stratification weights based on the population age structure in each study site. We capped the number of casual contacts reported at each location at 50, to be consistent with prior analyses and to reflect the low likelihood of increasing incremental risk of airborne transmission in settings with >50 people. We used the relative estimated incidence of tuberculosis by age group, sex and country derived from the 2023 Global Tuberculosis Report [1] to weight each contact by the risk that they had pulmonary tuberculosis. To account for differences in TB burden across demographic groups in exposure-hours calculation, these weights were normalized. (See Supplemental Methods for the full calculation)

For each country, we estimated the ‘exposure profile’ of participants by contact age and sex, representing the proportional risk of *Mtb* infection from different demographic subgroups. Exposure profiles combine information on close and causal contacts reported by participants and accounted for the risk of encountering a person with infectious tuberculosis by weighting each contact by the probability that they had pulmonary tuberculosis using country-, age-, and sex-specific tuberculosis incidence rates. We calculated assortativity (‘Q’) indices to quantify ‘like with like’ exposure-hours (e.g., women with women, 10–14-year-olds with other 10–14-year-olds) by age and sex for all contacts (close and casual combined).

The scripts and data for this analysis are available at: https://github.com/lopmanlab/GlobalMix-TB-analysis.

## Results

Participant characteristics are reported in detail elsewhere. Briefly, we recruited a total of 5,085 participants, an average of 1,271 per country. (Supplemental Table 1) All 10-year age bins (up to 60+) were represented, though we preferentially recruited children under 5 (28-30 % of the participants in each country), to ensure adequate representation of contact patterns in this group. Guatemala, in which the participants were 64% female, was the only country in which the female:male ratio was not nearly equal. The proportion of respondents currently in school was the lowest in Pakistan (15%) and highest in India (30%). Household sizes ranged from a median of 3 (IQR: 2, 5) in Guatemala to 6 (IQR: 4, 8) in Pakistan. (Supplemental Table 1)

Across the four countries, a total of 84,829 close contacts (14,707 in Guatemala, 20,270 in India, 17,674 in Mozambique, 32,178 in Pakistan) and 259,866 casual contacts (53,109 in Guatemala, 60,196 in India, 62,595 in Mozambique, and 83,966 in Pakistan) were reported. Close and casual contacts peaked among 10–19-year-olds in most countries, except in Pakistan, where they peaked among 5–9-year-olds. The number of close contacts tended to be similar among men and women except in Pakistan, where they were higher among men (Guatemala: p = 0.92, India: p = 0.11, Mozambique: p = 0.15, Pakistan: p < 0.05); casual contacts tended to be higher among men in all countries. (Supplemental Table 2)

We estimated daily exposure-hours (i.e., person-hours of possible *Mtb* exposure) from three sources: (1) contacts in the household, (2) non-household close contacts, (3) community locations in which participants spent time, in which they encountered casual contacts. Casual contacts made up the vast majority of exposure-hours reported in every country (42 exposure-hours in Guatemala and Mozambique, 66 exposure-hours in India, 76 exposure-hours in Pakistan). Exposure-hours to household (10 to 15 exposure-hours) and non-household close contacts (6 to 14 exposure-hours) were similar, but exposure-hours with household members exceeded exposure hours with non-household close contacts in all countries. (Figure 1)

**Figure 1.**
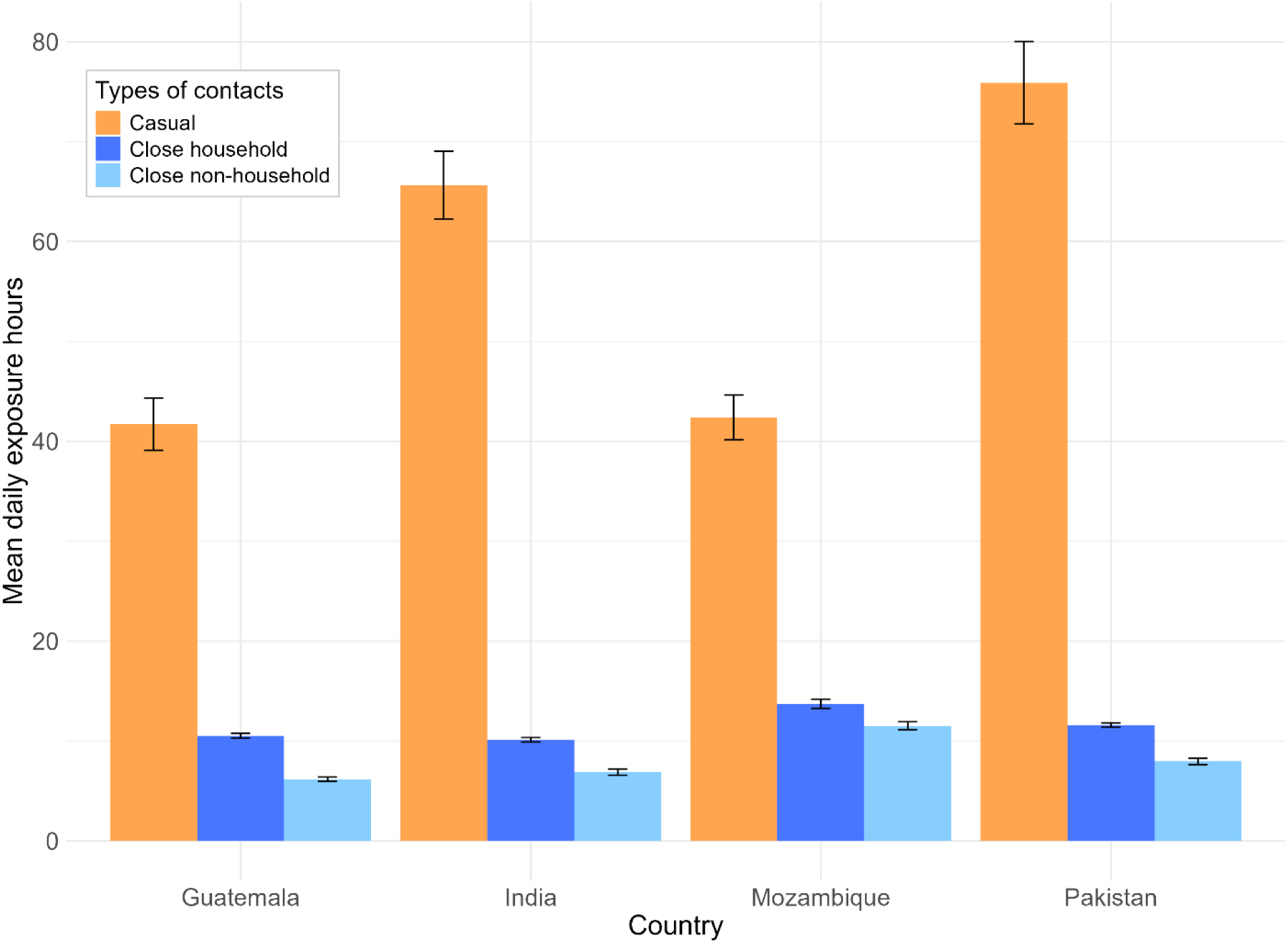
Mean daily exposure-hours by type of contact in four countries, 2021-2023 LEGEND: Mean daily exposure hours for three types of contacts, casual, close household, and close non-household contacts with 95% confidence intervals error bars. Exposure hours for casual contacts are calculated as contact time in hours multiplied by number of people present at each location of the visit. Exposure hours for close household and non-household contacts are calculated as the duration of contact in hours.

*Mtb* exposure profiles varied by age of the participant, though the trends were similar across countries. (Figure 2) The largest shifts in patterns of exposure by age occurred at the start of primary school (5-9 years) and early adulthood (20-29 years) in all countries. Age-assortativity indices were similar across countries (range:-0.078 – −0.066). Most exposure-hours occurred between men in all countries except for Guatemala, where exposure-hours were highest between women. The greatest sex-assortativity (men interacting with men and women with women) was in Pakistan (Q index: 0.3), a pattern which was present but less extreme in other countries (Q indices = 0.06-0.10). (Supplemental Figure 1) Children <5 years of age reported substantially more exposure-hours with women than with men in Mozambique and Guatemala (62% of exposure-hours in Guatemala were with women; 74% in Mozambique) but similar exposure-hours in Pakistan and India (50% of exposure-hours in Pakistan were with women; 54% in India).

**Figure 2.**
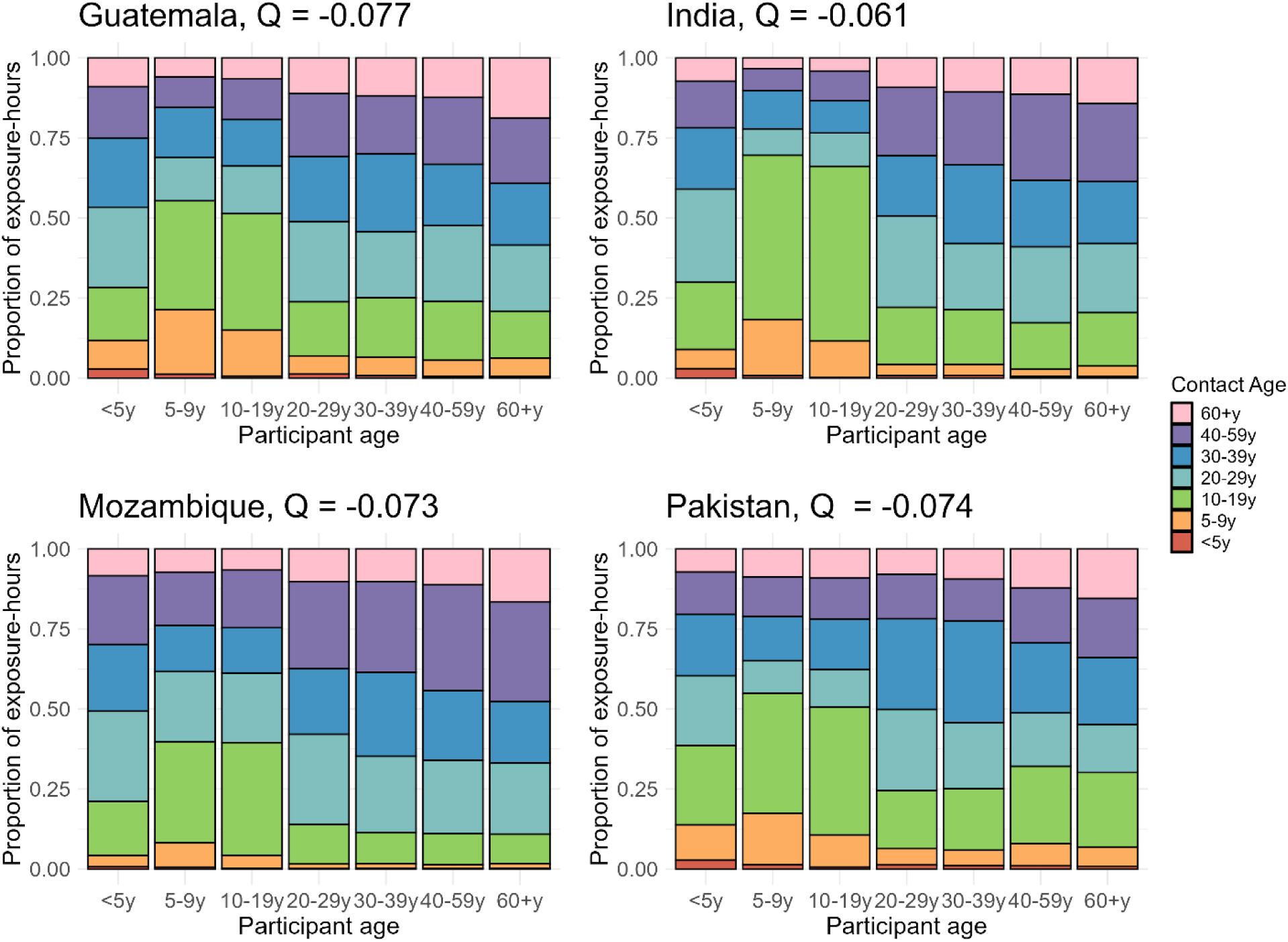
Exposure profiles by age of participants and their contacts in four countries, 2021-2023 LEGEND: Exposure-hours are weighted using population age structure and age-, sex-, and country-specific incidence of tuberculosis and calculated as duration of contacts multiplied by number of people present at each location summed across all locations for each age group. The Q index ranges from –1 to 1. Values near 1 indicate strong assortative mixing (participants preferentially interact with others of the same age group), values near –1 indicate disassortative mixing (participants preferentially interact with those from different age groups), and values near 0 suggest random mixing with respect to age.

To investigate the most likely community locations for *Mtb* exposure by country, we calculated the percentage of exposure-hours reported at non-home locations, weighting each contact by the likelihood they had pulmonary tuberculosis based on age-, sex-, and country-specific tuberculosis incidence estimates. There were differences in the locations most likely to be settings of *Mtb* transmission in each country. (Figure 3) In Guatemala and India overall, the most non-home exposure-hours were spent at school (29% in Guatemala and 41% in India). In Mozambique, the most non-home exposure-hours were reported at other peoples’ homes (26% of exposure-hours); in Pakistan, the most (22%) were reported on transit. As expected, the importance of locations varied by age. Among adults in Guatemala and older adults in Mozambique and Pakistan, places of worship made up 24-40% of non-home exposure hours. Other people’s homes were especially important places of exposure for children under 5 in all countries (21-54% of their non-home exposure-hours). However, the importance of school-based exposure varied: in India and Guatemala, 58-80% of exposure-hours among children 5-19 years were at school, as opposed to only 26-34%in Mozambique and Pakistan. In India and Guatemala, exposure-hours on transit were highest among older adults; in Pakistan, exposure-hours on transit were high across the age range. Exposure-hours at work in India were higher (greater than 30%) for adults, including those 60+, than in other countries. By comparison, exposure-hours at work were concentrated in a narrower age range in Guatemala, Mozambique, and Pakistan.

**Figure 3.**
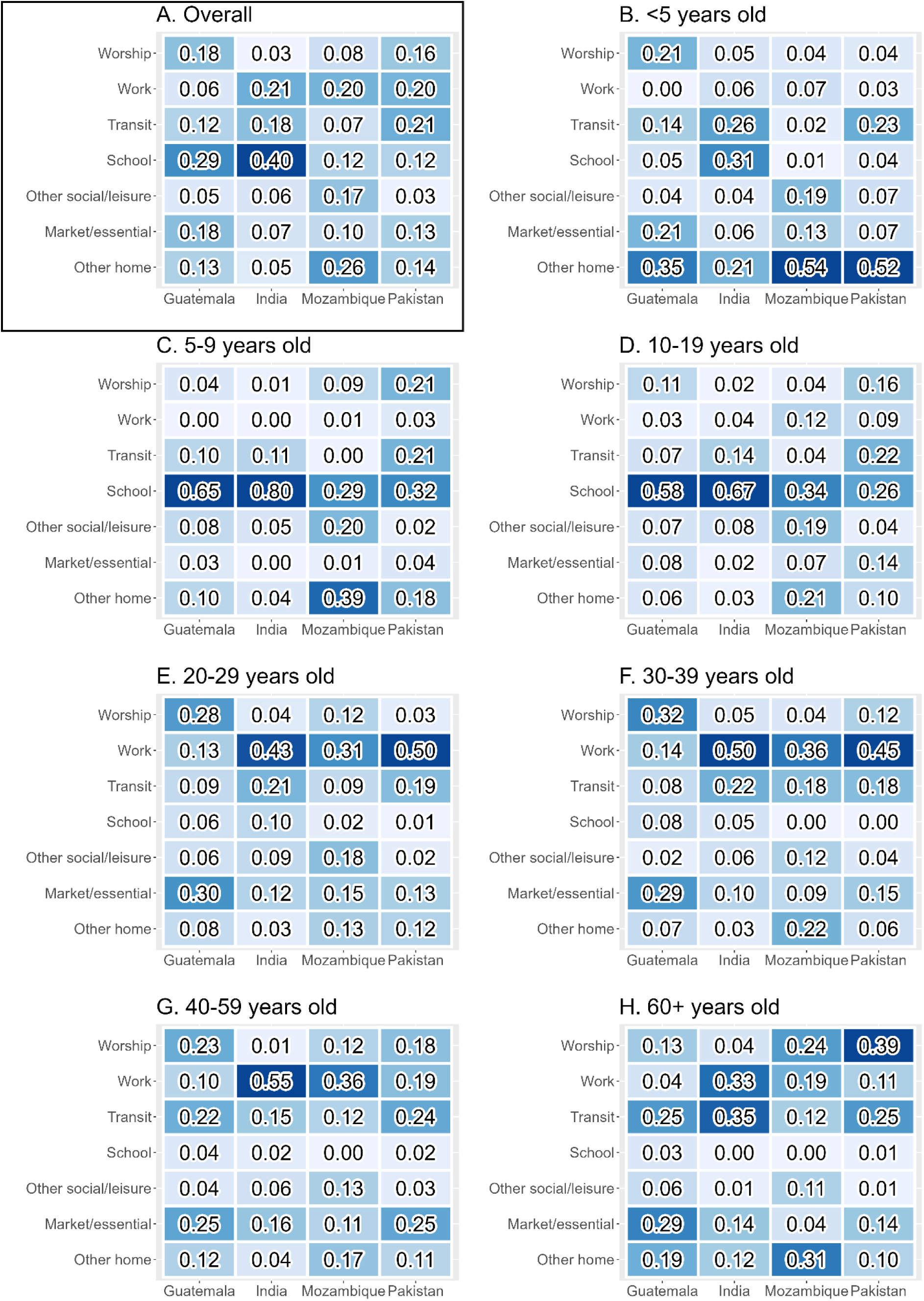
Proportion of exposure-hours associated with community locations in four countries by age and overall, 2021-2023 LEGEND: Values in each cell are the <u>proportion</u> of non-home exposure-hours reported at each location by country. Exposure-hours include those from both close and casual contacts and weighted by the point estimate of tuberculosis incidence in each country. Coloring of cells indicates fewer (light blue) to more (dark blue) exposure-hours reported in a location. The outlined box on the upper left shows exposure hours by location when summing across all age groups.

## Discussion

Using data on behavioral patterns from participants in four low- and middle-income countries, we characterized the relative importance of demographic groups and community locations in transmission of *Mtb*. In all countries, the majority of exposure-hours relevant for airborne transmission take place in non-home, community locations. The most prominent shifts in the profile of exposure occur at entry to primary school (5-9 years) and early adulthood (20-29 years). Schools, transit, markets, and places of worship varied in their relative importance as locations of transmission by country and age group. These findings underscore the importance of understanding local community structure and dynamics to optimally design interventions to prevent TB.

In high tuberculosis incidence settings, the relative importance of different types of contact in driving transmission has long been a source of controversy. Interventions like contact tracing that identify and screen close contacts are a mainstay of tuberculosis prevention programs because close, sustained contact is most likely to result in *Mtb* transmission; however, these interventions, when implemented at a population level, are not always effective in reducing overall tuberculosis incidence. [3], [21], [22] One reason for this may be that while close contacts are, on a per-capita basis, more likely to result in *Mtb* transmission than casual contacts, the greater number of casual contacts leads to their outsized importance in transmission overall. In this analysis, we found that 54-76% of exposure-hours occur among casual contacts. Notably, this proportion is well-aligned with the proportion of transmission events that cannot be explained by close contact in studies that combine genotyping (or genomic) and epidemiologic data to infer links between tuberculosis cases. [6], [7], [8], [9] When we compared the trends in contact to projections from the same countries used previously in tuberculosis transmission models, we found differences between country trends in contact by age and location. Namely, we found midlife increases in contact rates in some countries which was not accounted for in projections and a reduced importance of home locations in all countries, which is likely due to our capture of casual in addition to close contacts. (Supplemental Figure 3) Notably, previous projections showed much less variation across age groups in contact than our data and less variation overall between countries, underscoring the value of local data collection to describe context-specific contact patterns that may lead to *Mtb* transmission.

Age-sex patterns of exposure risk were similar across countries. School age children had the most dramatic shifts in exposure risk corresponding with the timing of typical school entry and exit. Protection from neonatal BCG vaccination wanes starting at age 5, increasing susceptibility to tuberculosis at the same time as primary school entry. [23] Simultaneously, immunological changes in early adolescence have been hypothesized to increase risk of progression to TB. [24] Disease type also shifts during this period, from the less transmissible paucibacillary disease characteristic of younger children to the more transmissible forms, including cavitary lung disease, typically seen in adults. [25], [26], [27], [28] Recognizing the social and behavioral shifts contributing to patterns of *Mtb* transmission – including dramatic changes in mobility and contact networks – as well as the biological ones, can help to understand how to best intervene to prevent tuberculosis in this age group. Children under 5 years old, a particularly important group given the challenges in diagnosing and treating tuberculosis at young ages, are mostly exposed to women. This is not surprising given the role that women play as primary caregivers of young children. Interventions that identify and screen pregnant women, young mothers and other caregivers could also be particularly important in reducing transmission to the youngest children. Finally, we saw substantial male-male assortativity in all countries except Guatemala, which is consistent with other findings that men drive transmission in high tuberculosis burden settings. [14] The relative importance of social and biological factors to this phenomenon is debated; our findings support the importance of behavioral differences in driving the persistent sex disparity in tuberculosis incidence.

Importantly, the type of community locations accounting for the most exposure-hours varied substantially by country. Schools made up 29% and 41% of all exposure-hours in Guatemala and India, underscoring the likely importance of schools in driving community-level *Mtb* transmission patterns in these settings. A study in South Africa, which incorporated CO_2_ measurements and contact data to infer the proportion of *Mtb* transmission events occurring in various community locations, estimated that more than 50% of transmission among 15-19 year-olds occurs in school settings. [19] Our results show that, the extent of *Mtb* transmission expected to take place in schools is large, underscoring the potential for school-based interventions to reduce overall community tuberculosis incidence. Public transit has previously been proposed as a setting that is conducive to transmission. However, we found that exposure-hours on transit varies substantially, from 8% of non-home exposure-hours in Mozambique to 22% of non-home exposure-hours in Pakistan. Therefore, interventions focused on transit may have varying effectiveness based on the setting. In Mozambique, others’ homes were a major source of contact exposure-hours (26%). Interactions in another person’s home are likely to rise to the level of ‘close’ contact, so it is likely that interventions that focus on identification of close contacts, such as contact tracing, would be particularly effective in these settings. On the contrary, contact tracing that is exclusively focused on household members of an index tuberculosis case may miss these important non-home exposures. Variation in the importance of locations by age can point towards useful avenues for intervention. Adults, who are responsible for the majority of *Mtb* transmission, reported a moderate number of exposure-hours in places of worship in most countries. (Indeed places of worship were notably locations of mass transmission events during the early COVID-19 pandemic. [29]) Though not often the setting of health interventions, places of worship may be excellent candidates for simple, inexpensive environmental changes such as increasing ventilation that could significantly reduce *Mtb* transmission.

There are limitations of our analysis. First, we collected data in Mozambique and Guatemala in 2021 and early 2022, during which time the COVID-19 pandemic could still have been affecting participants’ daily activities. However, despite changes in local policies that occurred during these periods, the number of contacts reported was relatively stable through multiple months of data collection in each site which suggests that the pandemic and associated policies were no longer severely affecting community activities. Second, we collected contact data from healthy community members. Participants may not represent individuals at highest risk for or with TB, or who currently have infectious TB, who may have systematically different contact patterns compared to healthy individuals. However, one benefit of data from healthy persons is that it is free from the reporting bias that can affect retrospective studies that ask tuberculosis cases to recall past contacts. Third, we set the upper limit for duration of exposure to a contact or a location as four hours, which was the longest option given in our survey. Some studies have shown gradations in *Mtb* infection among household contacts, suggesting fine differences in infection risk above four hours. [30], [31] However, data from outbreak investigations suggest that the threshold for an effective contact, or a contact which can result in *Mtb* transmission, is less than 10 hours. [32], [33] Our measure of exposure-hours accounts for the reported duration of the interaction, to acknowledge that duration of contact is likely an important modulating factor of transmission risk. Finally, we did not recruit across the entire year, so could not capture seasonal variations in contact patterns that may be important for transmission (i.e., holidays, school start and end, timing of annual migrations for employment). A previous study in India found higher rates of contacts in the winter season relative to the monsoon or summer seasons but these differences were slight. [34]

## Conclusions

Overall, our findings demonstrate consistency in age-specific contact patterns likely responsible for *Mtb* transmission across settings. Our data also support the notion that casual contacts in the community could be responsible for a large share of *Mtb* transmission. We also find that the likely locations of community transmission are highly varied across settings, underscoring the importance of interventions designed specifically for the communities in which they will be implemented.

## Data Availability

All data produced are available online at https://github.com/lopmanlab/GlobalMix-TB-analysis

## List of abbreviations

TB: Tuberculosis
Mtb: Mycobacterium tuberculosis

## Declarations

### Ethics approval and consent to participate

Informed consent was obtained from all participants 18 years old or older to participate in the study. For all individuals younger than 18 years, informed consent was obtained from their parents and guardians for their children and assent was obtained from individuals 12-17 years old. Consent of household heads was obtained for household participation. This study was conducted in accordance with the Declaration of Helsinki, and was approved by Emory University Institutional Review Board (approval number 00105630), Yale University (reliance agreement approval number 2000026911), UT Southwestern (reliance agreement approval number STU-2023-1107), and review boards at Christian Medical College Vellore (Approval #12065), Universidad del Valle de Guatemala (Protocol #223-12-2020) and the National Ethics Committee from the Ministry of Public Health and Social Assistance in Guatemala (Protocol #31-2020), Manhiça Health Research Institute Internal Scientific Committee (CCI/03/2020) and Internal Ethical Review Board (initial approval CIBS-CISM/011/2020), and The Aga Khan University (Approval #2022-7912-23648).

### Consent for publication

Not applicable

### Availability of data and materials

The datasets generated and/or analyzed during the current study are available in the GlobalMix-TB-analysis repository, https://github.com/lopmanlab/GlobalMix-TB-analysis

### Authors’ contributions

KNN conceptualized the analysis and methodology and wrote the initial manuscript draft, MS and SB conducted data analysis, MS curated data, RS, TSB, LM, and CD provided critical feedback on manuscript drafts, and VR, CS, MK, HM, SO, and BL obtained funding for the GlobalMix study and led data collection efforts.

## Acknowledgements

Not applicable

