## Supplemental materials for "Characterizing social behavior relevant for tuberculosis transmission in four low- and middle-income countries"

Supplemental Methods: Calculation of weights to account for TB status by age, sex, and country in exposure-hours calculations

The weight is calculated as the number of estimated TB cases per age-sex stratum divided by the total number of estimated TB cases in the country.

Let:

- $I_{a,s}$ be the estimated number of TB cases taken for age group $a$ and sex $s$ from Global Tuberculosis Report [1],
- $N$ be the total number of TB cases across all age-sex groups (i.e. $N = \sum_{a,s} I_{a,s}$),

Then, the weight $w_{a,s}$ is calculated as:

$$w_{a,s} = \frac{I_{a,s}}{N}$$

To apply these weights for exposure-hour calculations, we normalized each weight by dividing it by the mean weight across all age-sex strata in the country. This maintains the relative difference in TB burden while centering the weights around 1, allowing them to be multiplied with exposure-hours to reflect the likelihood of contacts having TB.

Let:

- $w_{a,s}$ be the unnormalized weight for age group $a$ and sex $s$ calculated previously.
- $K$ be the number of age-sex strata in the country,

Then, the normalized weight $w^*_{a,s}$ is calculated as:

$$w^*_{a,s} = \frac{w_{a,s}}{\frac{1}{K}\sum_{a,s} w_{a,s}}$$

### Supplemental Results: Figures and Tables

#### Supplemental Table 1. Characteristics of study participants by country

| | Guatemala | | India | | Mozambique | | Pakistan | |
|---|---|---|---|---|---|---|---|---|
| N | 1141 | | 1244 | | 1364 | | 1337 | |
| | **n** | **%** | **n** | **%** | **n** | **%** | **n** | **%** |
| Age | | | | | | | | |
| <5y | 332 | 29.1% | 348 | 28.0% | 409 | 30.0% | 391 | 29.2% |
| 5-9y | 202 | 10.3% | 126 | 10.1% | 122 | 9.0% | 129 | 9.7% |
| 10-19y | 125 | 17.7% | 259 | 20.8% | 249 | 18.3% | 258 | 19.3% |
| 20-29y | 109 | 11.0% | 125 | 10.0% | 125 | 9.2% | 129 | 9.7% |
| 30-39y | 125 | 9.6% | 126 | 10.1% | 125 | 9.2% | 137 | 10.2% |
| 40-59y | 117 | 11.0% | 135 | 10.9% | 209 | 15.3% | 146 | 10.9% |
| 60+y | 131 | 11.5% | 125 | 10.0% | 124 | 9.1% | 133 | 10.0% |
| NA | 0 | 0.0% | 0 | 0.0% | 0 | 0.0% | 14 | 1.1% |
| Sex | | | | | | | | |
| Female | 735 | 64.4% | 643 | 51.7% | 666 | 48.9% | 656 | 49.1% |
| Male | 405 | 35.5% | 601 | 48.3% | 696 | 51.1% | 674 | 50.4% |
| NA | 1 | 0.9% | 0 | 0.0% | 1 | 0.0% | 7 | 0.5% |
| Highest education level attained* | | | | | | | | |
| Currently enrolled in school | 266 | 32.9% | 339 | 37.8% | 368 | 38.6% | 189 | 20.3% |
| None | 246 | 30.4% | 88 | 9.8% | 79 | 8.3% | 435 | 46.7% |
| Primary school | 163 | 20.1% | 82 | 9.2% | 293 | 30.7% | 121 | 13.0% |
| Secondary school | 67 | 8.3% | 292 | 32.6% | 163 | 17.1% | 181 | 19.4% |
| Some college / higher education | 22 | 2.7% | 78 | 8.7% | 8 | 0.8% | 5 | 0.5% |
| NA | 45 | 5.6% | 17 | 1.9% | 43 | 4.5% | 1 | 0.1% |
| Occupation** | | | | | | | | |
| Student | 190 | 27.5% | 233 | 30.3% | 247 | 29.7% | 115 | 14.1% |
| Semiprofessional / professional | 28 | 4.1% | 32 | 4.2% | 83 | 10.0% | 54 | 6.6% |
| Skilled / semiskilled | 134 | 19.4% | 155 | 20.1% | 216 | 26.0% | 222 | 27.2% |
| Unemployed outside home | 332 | 48.0% | 274 | 35.6% | 214 | 25.7% | 392 | 48.0% |
| Other | 8 | 1.2% | 76 | 9.9% | 72 | 8.7% | 34 | 4.1% |
| Household size (categories) | | | | | | | | |
| 0-2 | 293 | 25.7% | 160 | 12.9% | 126 | 9.2% | 35 | 2.6% |
| 3-5 | 704 | 61.7% | 776 | 62.4% | 550 | 40.4% | 570 | 42.6% |
| 6-10 | 139 | 12.2% | 297 | 23.9% | 510 | 37.4% | 606 | 45.3% |
| 11+ | 4 | 0.4% | 11 | 0.9% | 63 | 4.6% | 126 | 9.4% |
| NA | 1 | 0.9% | 0 | 0.0% | 114 | 8.4% | 0 | 0.0% |

| | | | | | | | | |
|---|---|---|---|---|---|---|---|---|
| Median household size (IQR) | 2 (3, 5) | | 4.5 (4, 5) | | 5 (4, 7) | | 6 (4, 8) | |
| Generations in household | | | | | | | | |
| 1 | 247 | 21.6% | 148 | 11.9% | 303 | 22.2% | 39 | 2.9% |
| 2 | 827 | 72.5% | 673 | 54.1% | 630 | 46.1% | 1128 | 84.3% |
| 3 | 67 | 5.9% | 423 | 34.0% | 419 | 30.7% | 168 | 12.6% |
| NA | 0 | 0.0% | 0 | 0.0% | 12 | 0.9% | 2 | 0.1% |
| Multi-family household | | | | | | | | |
| No | 1024 | 89.7% | 1018 | 81.8% | 993 | 72.8% | 1217 | 91.0% |
| Yes | 117 | 10.3% | 226 | 18.2% | 359 | 26.3% | 118 | 8.8 |
| NA | 0.0 | 0.0% | 0 | 0.0% | 12 | 0.9% | 2 | 0.1% |

* Among participants over 5 years of age; in Mozambique, missing category includes some 5- to 9-year-olds who have not yet entered school

** Among participants 10 years of age and older

### Supplemental Table 2. Contacts relevant for *Mtb* transmission in four countries, 2021-2023

| | **Guatemala** | | **India** | | **Mozambique** | | **Pakistan** | |
|---|---|---|---|---|---|---|---|---|
| **Overall, median (IQR)** | 19 (14, 43) | | 23 (13, 49) | | 26 (15, 48) | | 33 (18, 68) | |
| | **Close** | **Casual** | **Close** | **Casual** | **Close** | **Casual** | **Close** | **Casual** |
| | **Median (IQR)** | **Median (IQR)** | **Median (IQR)** | **Median (IQR)** | **Median (IQR)** | **Median (IQR)** | **Median (IQR)** | **Median (IQR)** |
| Overall | 7 (5, 7) | 12 (8, 35) | 7 (6, 10) | 15 (6, 38) | 6 (4, 9) | 17 (9, 37) | 11 (7, 17) | 21 (9, 50) |
| Participant age | | | | | | | | |
| <5y | 6 (6, 7) | 8 (6, 11) | 7 (6, 9) | 7 (5, 11) | 5 (3, 8) | 12 (8, 20) | 8 (6, 12) | 12 (6, 24) |
| 5-9y | 7 (6, 8) | 17 (8, 35.8) | 10 (7, 12) | 23 (8, 44.2) | 6 (4, 10) | 19 (9.5, 36) | 13.5 (9, 18) | 31 (15, 68) |
| 10-19y | 7 (6, 8) | 23 (10, 41) | 10 (8, 12) | 34 (14, 51.8) | 8 (5, 11) | 27 (14, 46) | 12 (9, 17) | 27.5 (12, 65.8) |
| 20-29y | 6 (5, 8) | 13 (7.8, 45) | 8 (6, 9) | 15.5 (7, 34) | 6 (4, 9) | 21 (10, 47) | 11 (7, 16) | 18 (9, 41) |
| 30-39y | 6 (5, 7) | 13 (8, 47.8) | 7 (5, 10) | 13 (5, 31.2) | 6 (4, 9) | 15 (8, 28) | 13 (9, 19) | 26 (10, 75) |
| 40-59y | 7 (6, 7) | 11 (8, 30) | 7 (5, 9) | 15 (7, 44) | 5 (3, 9) | 13 (7, 31.2) | 11 (7, 16) | 19 (9, 48) |
| 60+y | 6 (4, 7) | 9.5 (6, 20.8) | 6 (4, 8) | 10 (5, 25) | 5 (2.5, 7) | 8 (5, 15) | 12 (8, 17) | 17 (9, 30) |
| Participant sex | | | | | | | | |
| Female | 6 (5, 7) | 11 (7, 32) | 7 (6, 10) | 11 (5, 31) | 6 (3, 9) | 14 (7.8, 27) | 9 (6, 15) | 14 (6, 28) |

| | | | | | | | | |
|---|---|---|---|---|---|---|---|---|
| Male | 6 (6, 7) | 10 (7, 25) | 8 (6, 10) | 15 (7, 38) | 6 (4, 9) | 16 (9, 32) | 12 (8, 17) | 25 (12, 62) |

Supplemental Figure 1. Daily exposure-hours by sex of participant and contact

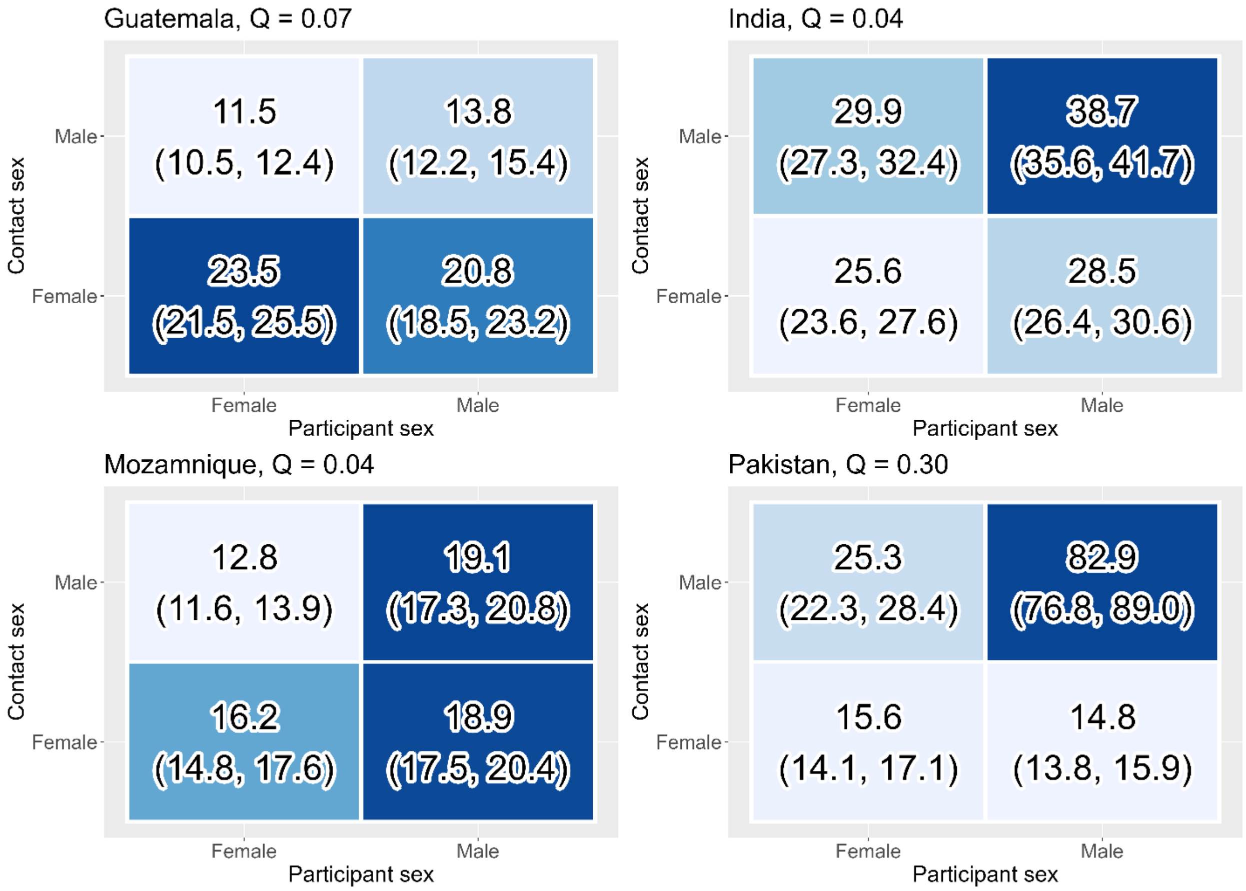


LEGEND: Exposure-hours are weighted by population age structure and tuberculosis incidence and calculated as duration of contacts multiplied by number of people present at each location summed across all locations for each sex category. Mean exposure-hours and 95% confidence intervals for each sex participant-contact combination is shown in a matrix form. The Q index ranges from –1 to 1. Values near 1 indicate strong assortative mixing (participants preferentially interact with others of the same age group), values near –1 indicate disassortative mixing (participants preferentially interact with those from different age groups), and values near 0 suggest random mixing with respect to age.

Supplemental Figure 2. Stringency of pandemic restrictions in place in each country during study period

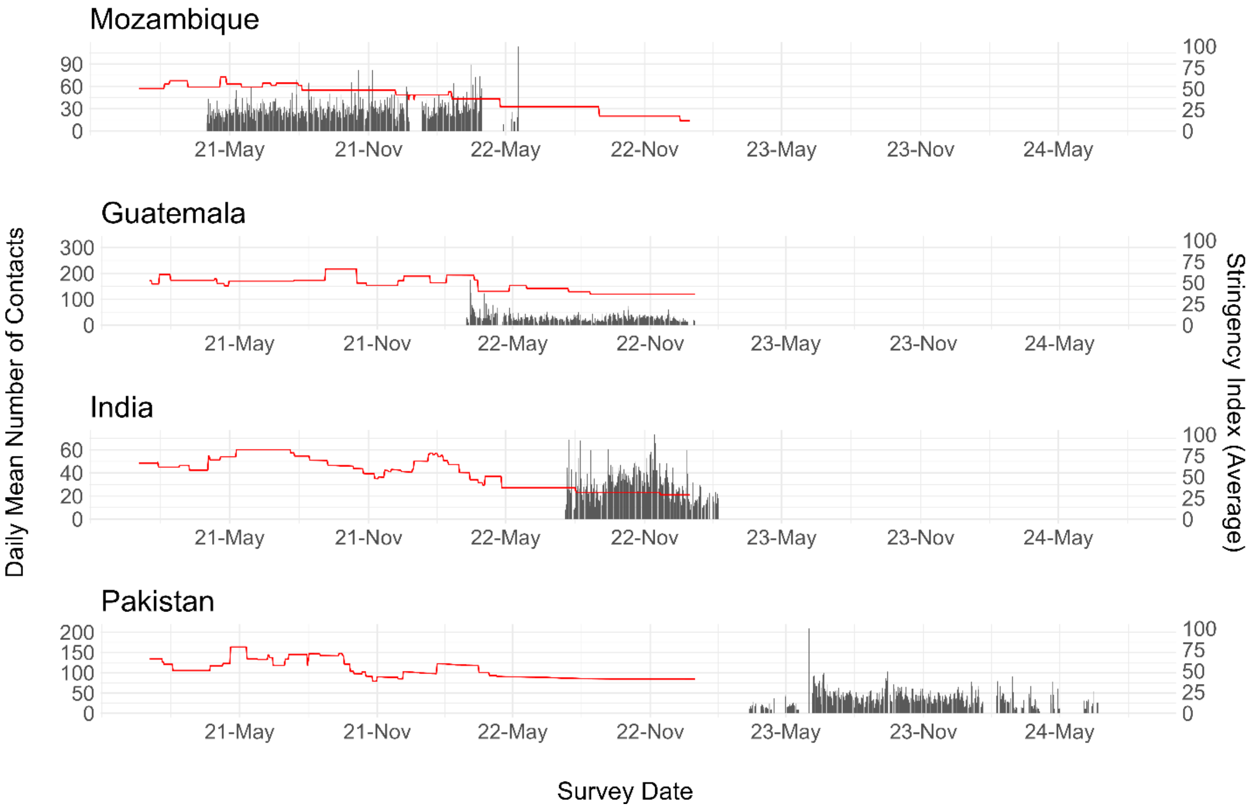


LEGEND: Daily mean number of contacts (including close and casual contacts) by dates are shown against the Oxford COVID-19 Government Response Tracker (OxCGRT) stringency index (in red) [1], which tracked pandemic-era government policies related to closure and containment of SARS-CoV-2 spread and provide a snapshot of the number and degree of policies in place in a given area.

Supplemental Figure 3. Comparison of contact rate estimates by age and location of contact with POLYMOD-based projections

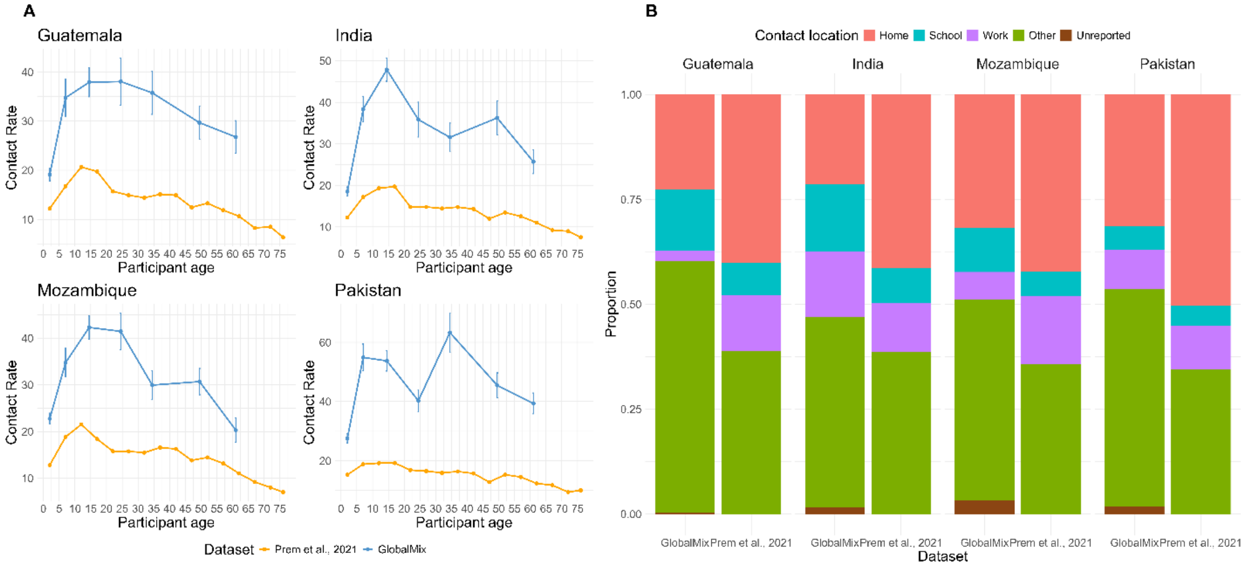


LEGEND: A. Mean contact rates by age estimated by Prem et al. [2] based on the POLYMOD data and the contact rates estimated in our study, GlobalMix (close and casual contact combined). 95% confidence intervals are shown with vertical bars.
B. proportions of contacts reported in each location, categorized using the scheme in POLYMOD. Contacts in GlobalMix for which the location was unreported are shown in purple.
